# Adherence to the EAT-Lancet Healthy Reference Diet in relation to Coronary Heart Disease, All-Cause Mortality Risk and Environmental Impact: Results from the EPIC-NL Cohort

**DOI:** 10.1101/2021.06.30.21259766

**Authors:** Chiara Colizzi, Marjolein C Harbers, Reina E Vellinga, WM Monique Verschuren, Jolanda MA Boer, Elisabeth HM Temme, Yvonne T van der Schouw

## Abstract

**Objectives:** To construct a diet-score measuring the level of adherence to the Healthy Reference Diet (HRD), to explore whether adherence to the HRD is associated with coronary heart disease (CHD), all-cause mortality risk, and to calculate its environmental impact.

**Design:** Prospective cohort study.

**Setting:** The Dutch contribution to the European Prospective Investigation into Cancer and Nutrition (EPIC-NL).

**Participants:** 37,349 adults (20-70y) without CHD at baseline.

**Main outcome measures:** Primary outcomes were incident CHD and all-cause mortality. Secondary outcomes were greenhouse gas emission (GHGE), land use, blue water use, freshwater eutrophication, marine eutrophication, and terrestrial acidification.

**Results:** During a median 15.3-year follow-up, 2,543 cases of CHD occurred, and 5,648 individuals died from all causes. The average HRD-score was 73 (SD=10). High adherence to the HRD was associated with a 15% lower risk of CHD (hazard ratio 0.85, 95% confidence interval 0.75 to 0.96), as well as a 17% lower risk of all-cause mortality (hazard ratio 0.83, 95% confidence interval 0.77 to 0.90) in multivariable-adjusted models. Better adherence to the HRD was associated with lower environmental impact from GHGE (β= -0.10 kg CO2-eq, 95% confidence interval -0.13 to -0.07), land use (β= -0.11 m^2^ per year, 95% confidence interval -0.12 to -0.09), freshwater eutrophication (β= -0.000002 kg P-eq, 95% confidence interval -0.000004 to -0.000001), marine eutrophication (β= -0.00035 kg N-eq, 95% confidence interval -0.00042 to -0.00029), and terrestrial acidification (β = -0.004 kg SO2-eq, 95% confidence interval -0.004 to -0.003), but with higher environmental impact from blue water use (β=0.044 m^3^, 95% confidence interval 0.043 to 0.045).

**Conclusion:** High adherence to the HRD was associated with lower risk of CHD and all-cause mortality. Additionally, increasing adherence to the HRD could lower some aspects of the environmental impact of diets, but attention is needed for the associated increase in blue water use.

## 1. Introduction

Diet has a profound impact on human health as well as the environment.^1^ According to the Global Burden of Disease Study 2017, 11 million deaths and 255 million DALYs can be attributed to high sodium intake, and low intake of whole grains and fruit across the world.^2^ Unhealthy diets are considered one of the main risk factors for the development of cardiovascular diseases.^3^ At the same time, current dietary practices are likely to exhaust our planet in the light of the expected growth of the world population.^1^ Food production practices account for up to 30% of global greenhouse-gas emissions (GHGE) and 70% of freshwater use^1^, most of which is intended for meat and dairy production.^4-6^ For these reasons, shifting towards healthy and sustainable diets could co-benefit public and planetary health.

The EAT-Lancet Commission on Healthy Diets From Sustainable Food Systems is the first large-scale and coordinated scientific collaboration to provide dietary guidelines on healthy diets within the food production boundaries for the world population.^7^ The commission proposed the Healthy Reference Diet (HRD), that was constructed based on scientifically established targets for healthy diets and fitting within a safe operating space of food systems, for which the Planetary Boundaries framework was used. The diet includes high consumption of fruits and vegetables, whole grains, legumes, nuts, and unsaturated oils; low to moderate consumption of dairy, starchy vegetables, poultry and fish; and no or low consumption of saturated fats, red meat, and all sweeteners.^7^ As such, the HRD generally emphasizes the intake of plant-based foods and suggests to limit the intake of animal-sourced foods and starchy vegetables.

There is still limited evidence directly linking the HRD to cardiovascular outcomes and mortality. The EAT-Lancet report projected that 19.0-23.6% of premature adult deaths could potentially be avoided by adopting the HRD, while remaining within acceptable environmental boundaries.^7^ However, these projections were based on theoretical models. To date, only one study empirically assessed the association between the HRD and the risk of coronary heart disease (CHD) and all-cause mortality, showing that better adherence to the HRD was associated with 28% lower risk for CHD, but not with risk of stroke or all-cause mortality.^8^ Potentially, this may relate to the dichotomous scoring system that was applied, which consequently did not allow for large variation in HRD-scores. Thus, evidence on the potential cardiovascular benefits of the HRD coming from prospective cohort studies using a refined diet-score to measure adherence is currently lacking. Additionally, the environmental impact of the HRD has not been previously assessed empirically. Insight into the cardiovascular and planetary consequences of adhering to the HRD would help to identify win-win or win-lose aspects of the HRD.

Therefore, the present study aimed to construct a refined HRD-score allowing for wide variation in adherence to the HRD. Second, we aimed to estimate the association of adherence to the HRD with CHD and all-cause mortality risk in a population-based cohort study. Third, we aimed to estimate the associated environmental impact of the HRD using a wide range of environmental indicators relating to the planetary boundaries in the same population-based cohort study.

## 2. Methodology

### Study population

We used data from the Dutch contribution to the European Prospective Investigation into Cancer and Nutrition (EPIC-NL).^9^ The EPIC study was designed to assess the associations between diet, lifestyle, dietary intake, and the incidence of cancer and other chronic conditions [9]. The EPIC-NL cohort combines the MORGEN cohort (*n*[=[22,654) and the Prospect cohort (*n* = 17,357), resulting in a total of 40,011 participants. The MORGEN cohort included both men and women, aged 20-64 years, from three Dutch cities (Amsterdam, Doetinchem, and Maastricht), recruited between 1993 and 1997. The Prospect cohort included women participating in a breast screening program, aged 49-70 years, recruited between 1993 and 1995 from Utrecht and its vicinity. At baseline, participants completed a general questionnaire and a validated semi-quantitative food frequency questionnaire (FFQ). During a physical examination a non-fasting blood sample was taken, aliquoted and stored for future research. The EPIC-NL study was conducted according to the guidelines in the Declaration of Helsinki and all procedures involving the participants were approved by the institutional review board of the University Medical Center Utrecht (Prospect-EPIC) and the medical ethical committee of TNO Nutrition and Food Research (MORGEN-EPIC). All participants provided written informed consent.

For the current study, we excluded participants who withheld permission for linkage with national disease registries (n=1,666), those who withdrew informed consent during follow-up (n=1), participants with prevalent CHD at baseline (n=377), participants with missing dietary intake data (n=218), and particpants with implausible energy intake (defined as those in the lowest and highest 0.5% of the ratio of energy intake over basal metabolic rate) (n=400), leaving 37,349 persons for analysis (Supplementary Figure 1).

### Calculation of the HRD adherence score

The FFQ included questions on the consumption of 178 food items in the year prior to enrolment.^9 10^ For some food items, questions were accompanied by images of the food in different portion sizes, to assist in portion size estimation. Frequency of consumption was estimated in times per day, week, month, year or never. Average food intake (g/d) was calculated by multiplying the consumption frequency with the consumed amounts and nutrient intakes were calculated using the Dutch food composition table of 1996.^11^

To asses adherence to the HRD, a Healthy Reference Diet score (HRD-score) was constructed. To calculate the adherence scores, the dietary recommendations from the EAT-Lancet report were recalculated on the basis of 2000 kcal/day for women, in line with the recommended energy intake proposed by the Dutch dietary guidelines (Supplementary Table 1 and 2). Participants were assigned proportional scores ranging from 0-10 for each of the 14 dietary recommendations in the HRD (as proposed by EAT-Lancet), that were then summed, resulting in a score ranging between 0 (no adherence) and 140 (complete adherence). Each food group in the HRD-score was categorized into one of the following scoring components adapted from Looman et al.^12^: adequacy, moderation, optimum or ratio. The allocation of scoring components to the dietary recommendations in the HRD was informed by literature investigating the associations of those food groups with chronic disease.^13-35^ Adequacy components are used to score foods generally considered healthy and for which a high intake is recommended. In the HRD-score, foods assigned to this component were whole grains, vegetables, fruits, legumes, and soy foods. Participants received 10 points for meeting the recommended intake for these food groups, 0 points for no consumption, and a proportional score for intakes between zero and the recommended level. Moderation components were used to score foods that could increase the risk of chronic diseases. The moderation component was used to score beef, lamb, pork, and sweeteners. For these foods, 0 points were assigned if the intake was above the reference intake, 10 points were assigned for an intake equal to or lower than the reference intake, and intermediate intakes were assigned a proportional scoring.

**Table 1.** Baseline characteristics of the EPIC-NL cohort by quartiles of the HRDea-score (*n*=37,349)^1^

|  | Quartiles of HRDea-scores (range) |  |  |  |
| --- | --- | --- | --- | --- |
|  | Q1 (32-66)<br>N=9340 | Q2 (67-73)<br>N=9340 | Q3 (74-79)<br>N=9340 | Q4 (80-116)<br>N=9340 |
| Sex |  |  |  |  |
| Male | 3773 (40.4) | 2753 (29.5) | 1799 (19.3) | 1086 (11.6) |
| Female | 5567 (59.6) | 6587 (70.5) | 7541 (80.7) | 8254 (88.4) |
| Age | 48.5 (36.8, 55.5) | 51.0 (41.4, 57.3) | 52.5 (44.9, 58.6) | 52.7 (46.9, 58.9) |
| BMI |  |  |  |  |
| Normal weight | 4114 (44.7) | 4391 (47.7) | 4359 (47.4) | 4848 (52.7) |
| Overweight | 3731 (40.5) | 3559 (38.6) | 3640 (39.6) | 3319 (36.1) |
| Obesity | 1357 (14.7) | 1261 (13.7) | 1195 (13.0) | 1035 (11.2) |
| Educational level |  |  |  |  |
| Low | 5912 (63.7) | 5612 (60.4) | 5375 (57.8) | 4639 (49.9) |
| Moderate | 2122 (22.9) | 2036 (21.9) | 1951 (21.0) | 1976 (21.3) |
| High | 1250 (13.5) | 1645 (17.7) | 1966 (21.2) | 2681 (28.8) |
| Smoking |  |  |  |  |
| Never | 3152 (33.9) | 3487 (37.5) | 3751 (40.3) | 3852 (41.3) |
| Former | 2466 (26.5) | 2860 (30.7) | 3151 (33.9) | 3397 (36.5) |
| Current | 3687 (39.6) | 2958 (31.8) | 2403 (25.8) | 2067 (22.2) |
| Physical activity |  |  |  |  |
| Inactive | 889 (9.5) | 728 (7.8) | 666 (7.1) | 524 (5.6) |
| Moderately inactive | 2259 (24.2) | 2371 (25.4) | 2348 (25.1) | 2294 (24.6) |
| Moderately active | 2260 (24.2) | 2412 (25.8) | 2442 (26.1) | 2588 (27.7) |
| Active | 3932 (42.1) | 3829 (41.0) | 3884 (41.6) | 3934 (42.1) |
| Alcohol consumption, g/day | 5.3 (0.7, 17.3) | 4.9 (0.7, 15.6) | 4.8 (0.7, 14.8) | 5.0 (0.7, 14.8) |
| Energy intake, kcal/day | 2269 (1872, 2738) | 2020 (1704, 2424) | 1860 (1568, 2199) | 1736 (1473, 2046) |
| Food consumption, g/day |  |  |  |  |
| Whole grains | 20.7 (2.4, 85.2) | 49.1 (6.4, 107.4) | 72.1 (15.0, 127.4) | 96.5 (46.8, 134.1) |
| Vegetables | 89.5 (66.7, 117.0) | 99.2 (74.7, 129.4) | 105.9 (81.9, 136.9) | 125.8 (95.4, 166.2) |
| Fruit | 105.1 (49.8, 180.7) | 136.9 (90.7, 250.2) | 190.6 (122.4, 278.7) | 241.4 (158.4, 323.5) |
| Potatoes and cassava | 142.9 (105.1, 183.9) | 106.8 (67.9, 156.9) | 76.9 (49.6, 111.4) | 60.1 (38.7, 81.0) |
| Dairy foods<sup>2</sup> | 538.6 (258.5, 722.6) | 417.5 (232.4, 613.2) | 381.0 (232.5, 543.5) | 323.6 (207.8, 427.2) |
| Legumes<sup>3</sup> | 23.7 (14.7, 35.9) | 26.9 (17.2, 40.2) | 29.0 (18.8, 42.0) | 33.8 (23.2, 46.4) |
| Soy | 0.0 (0.0, 0.0) | 0.0 (0.0, 0.0) | 0.0 (0.0, 0.1) | 0.0 (0.0, 4.8) |
| Beef, lamb, and pork | 103.7 (70.8, 136.3) | 93.5 (59.3, 123.9) | 82.8 (51.9, 111.4) | 64.0 (34.5, 97.5) |
| Chicken | 10.3 (4.7, 17.3) | 9.6 (4.3, 16.3) | 9.4 (4.1, 15.9) | 7.9 (2.9, 14.6) |
| Eggs | 21.4 (10.5, 28.6) | 14.3 (7.6, 21.4) | 13.8 (7.1, 17.6) | 10.5 (5.8, 15.7) |
| Fish | 7.2 (2.8, 14.0) | 7.4 (3.3, 14.0) | 8.0 (3.3, 15.2) | 8.4 (3.3, 15.9) |
| Nuts | 4.3 (1.4, 11.7) | 4.1 (1.4, 10.7) | 3.8 (1.4, 9.7) | 4.7 (1.7, 11.4) |
| Unsaturated fats | 11.3 (4.9, 20.8) | 10.9 (5.1, 19.6) | 10.1 (5.0, 17.8) | 9.9 (5.1, 16.9) |
| Saturated fats | 33.5 (21.4, 49.2) | 28.5 (17.6, 42.5) | 24.7 (14.4, 37.8) | 22.3 (12.3, 35.0) |
| Added sugars | 193.7 (117.4, 305.9) | 178.4 (108.9, 271.4) | 170.1 (101.3, 251.0) | 159.8 (93.3, 231.7) |

<sup>1</sup>Estimates are presented as counts n and percentages (%) or as medians (p25, p75). <sup>2</sup>Including whole milk,
derivate equivalents and cheese. <sup>3</sup>Including beans, lentils, and peas.

**Table 2.**
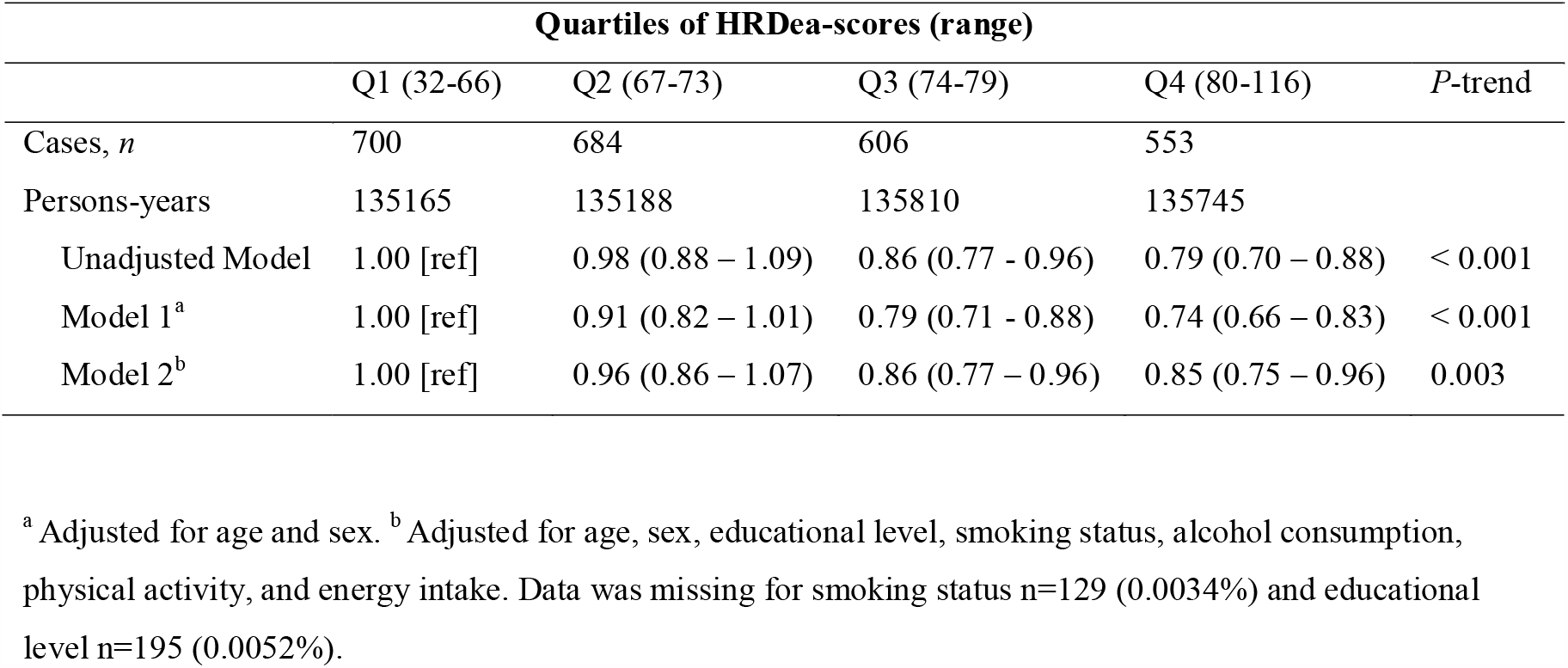
Hazard ratios (HRs) and 95% confidence intervals (CIs) for the association between quartiles of the HRDea-score and incident of CHD (*n*=37,349).

|  | Quartiles of HRDea-scores (range) |  |  |  | P-trend |
| --- | --- | --- | --- | --- | --- |
|  | Q1 (32-66) | Q2 (67-73) | Q3 (74-79) | Q4 (80-116) |  |
| Cases, *n* | 700 | 684 | 606 | 553 |  |
| Persons-years | 135165 | 135188 | 135810 | 135745 |  |
| Unadjusted Model | 1.00 [ref] | 0.98 (0.88 – 1.09) | 0.86 (0.77 - 0.96) | 0.79 (0.70 – 0.88) | < 0.001 |
| Model 1<sup>a</sup> | 1.00 [ref] | 0.91 (0.82 – 1.01) | 0.79 (0.71 - 0.88) | 0.74 (0.66 – 0.83) | < 0.001 |
| Model 2<sup>b</sup> | 1.00 [ref] | 0.96 (0.86 – 1.07) | 0.86 (0.77 – 0.96) | 0.85 (0.75 – 0.96) | 0.003 |

<sup>a</sup> Adjusted for age and sex. <sup>b</sup> Adjusted for age, sex, educational level, smoking status, alcohol consumption, physical activity, and energy intake. Data was missing for smoking status *n*=129 (0.0034%) and educational level *n*=195 (0.0052%).

Optimum components comprise foods which are nutritious yet potentially detrimental if eaten in large quantities on a daily basis. The optimum component was used to score the following food groups: potatoes, dairy, chicken, eggs, fish, and nuts. For these foods, participants with intakes within the required optimum intake range would receive 10 points, while those with intakes lower or higher than the optimum would be scored proportionally and symmetrically from 0 to 10 and from 10 to 0. Finally, a ratio component was used to describe the added fats food group. For the added fats, no consumption of unsaturated fats or an unsaturated to saturated fats ratio lower than 0.6 was assigned 0 points, while no consumption of saturated fats or an unsaturated to saturated fats ratio higher than 13 was assigned 10 points. Ratios in between were scored proportionally. Cut-offs and threshold values for the ratio component were derived from the 15th percentile and 85th percentile of the intake distribution of the Dutch reference population, as described in Looman et al.^12^

Finally, the HRD-score was adjusted for energy intake (HRDea-score) using the energy-adjusted nutrient residual model to remove the variance in dietary intake related to total energy intake.^36^

### CHD and all-cause mortality ascertainment

CHD events included both fatal and non-fatal cases of CHD. Morbidity data were obtained from the Dutch Center for Health Care Information, which holds a standardized computerized registry of hospital discharge diagnoses. The hospital discharge diagnosis database was linked to the cohort based on information of birthdate, sex, postal code, and general practitioner with a validated probabilistic method.^37^ Hospitalization for CHD was based on the principal diagnoses (ICD 10: I20-I25).

Information on vital status was obtained through linkage with the Dutch municipal registry. All-cause mortality was defined as death from any cause after study inclusion. For deceased participants, information on the causes of death was ascertained through linkage with the Causes of Death Registry of Statistics Netherlands. Death from CHD was based on both primary and secondary causes of death. A primary cause of death was defined as death due to a CHD event, while a secondary cause of death was defined as death due to complications of the primary cause, or another disease which could have led to death. All participants were followed until CHD event, death, emigration, or end of follow-up, whichever came first. Follow-up was complete until December 31st, 2010.

### Environmental impact assessment

The ‘planetary boundaries’ within the planetary boundaries framework provide the safe operating space for the Earth’s biophysical subsystems and or processes,^38^ and also underlie the EAT-Lancet’s commission’s environmental impact assessments. Within the planetary boundaries framework, the main environmental systems and processes that are affected by food production are climate change, biodiversity loss, land system change, freshwater use, and nitrogen and phosphorus flows.^7^ Within this framework, the state of these systems is further defined by so-called control variables. As the main environmental systems are interlinked and interdependent, most control variables relate to multiple environmental systems. For example, greenhouse gas emissions (GHGE) are an indicator of biodiversity loss and climate change; land use is an indicator of biodiversity loss and land system change; blue water use (e.g, irrigation water) is an indicator of biodiversity loss and freshwater use; eutrophication (e.g., through application of fertilizer) is an indicator of nitrogen and phosphorus cycles, biodiversity loss and climate change, and terrestrial acidification is an indicator of biodiversity loss.^7 38 39^ Therefore, the assessment of a wide range of environmental indicators provides a holistic assessment of the environmental impact of the HRD. In the present study, we evaluate the effects of the HRD on GHGE (kg CO_2_-eq per day), land use (m^2^ per year), blue water use (m^3^ per day), freshwater eutrophication (kg P-eq per day), marine eutrophication (kg N-eq per day), and terrestrial acidification (kg SO2-eq day).

The associated environmental impact of the 178 foods and beverages were assessed using the most recent Life Cycle Assessments (LCA) data from the Dutch LCA Food database.^40^ This database is established by the National Institute for Public Health and the Environment (RIVM) and contains information on the environmental impact for approximately 250 Dutch foods and beverages. A full description of the data and assumptions can be found elsewhere.^41^ In short, the LCAs had an attributional approach and hierarchical perspective. System boundaries were from cradle till plate, including primary production, processing, primary packaging, distribution, retail, supermarket, storage, preparation by the consumer (e.g., cooking), and incineration of packaging waste. Transport between all phases, except from retail to the consumer was included. Economic allocation was applied for all food items, except for milk, where physical allocation was used. In order to estimate daily environmental impact, LCA data from the Dutch LCA Food database, referred to as primary data, was linked via NEVO-codes to FFQ items. Extrapolations were carried out in case no primary LCA data were available.

### Ascertainment of covariates

Details on data collection on covariates are described elsewhere.^9^ In short, for age, sex, educational level, smoking status and history, physical activity, and medication use data from the baseline general questionnaire were used. Education was categorized into low (lower vocational training and primary school), moderate (secondary school and intermediate vocational training), and high educational level (higher vocational training and university). Smoking status was categorized into never smoker, former smoker, or current smoker. Alcohol intake was assessed from the FFQ, and measured in grams/day. Physical activity was categorized into inactive, moderately inactive, moderately active, and active, according to the Cambridge Physical Activity Index (CPAI).^42^ Total energy intake was also derived from the FFQ, and expressed in kilocalories/day.

The baseline physical examination provided data on body weight and height, blood pressure and cholesterol levels [10]. BMI was calculated as height divided by weight squared, and participants were categorized as normal weight for a BMI ≤ 24.9 kg/m^2^, overweight for a BMI between 25 and 29.9 kg/m^2^, and obese for a BMI ≥ 30 kg/m^2^. Both systolic and diastolic blood pressure were measured twice in supine position, from which the mean was taken. Blood pressure measurements were performed on the left arm, using a Boso Oscillomat in the MORGEN-EPIC cohort, and a random zero Sphygmomanometer in the Prospect-EPIC cohort.^9^ Hypertension was defined as use of hypertensive medication, and/or systolic blood pressure >140 and/or diastolic pressure >90.^9^ Serum total cholesterol (mmol/l) was measured using enzymatic methods.^9^

### Statistical analysis

All baseline characteristics are reported by quartiles of the HRDea-score. Normally distributed continuous variables are presented as means with standard deviations. Continuous variables with a skewed distribution are presented as median with interquartile range (IQR). Categorical variables are presented as counts and percentages. A Cox proportional hazard model was used to obtain hazard ratios (HR) and 95% confidence intervals (CI) for the association between quartiles of the HRDea-score and CHD risk and all-cause mortality. The lowest quartile was used as reference. The underlying time variable was age from study entry to either diagnosis, death, or end of follow-up (31-12-2010), whichever came first. The proportional hazards assumption was checked using the Schoenfeld test, with no violations observed.

For CHD and all-cause mortality outcomes, the analyses present first the unadjusted model with crude estimates. Model 1 was adjusted for age and sex and model 2 was additionally adjusted for educational level, smoking, alcohol consumption, physical activity, and energy intake. A sensitivity analysis was conducted, where we added BMI, total cholesterol, and hypertension to the multivariable-adjusted model (model 2), as these factors may be potential mediators in the association between the HRDea-score and CHD. All mediators were first added individually and then simultaneously.

All foods in the FFQ, expressed in grams/day, had an estimated environmental impact calculated with LCA. We used linear regression models to estimate the association between HRDea-score and each environmental indicator. In this linear regression the exposure was the HRDea-score and the outcome was the environmental indicator, calculated as the sum of the associated environmental impact of the food groups included in the HRD. The lowest quartile was used as reference. The analyses present first the crude estimates, and then in model 1 estimates were adjusted for age, sex, and energy intake. No other variables were included, based on current literature on the environmental impact of diets. The p-value for trend across quartiles was estimated by modelling the median value of each quartile as a continuous variable. Statistical significance was set at a two-tailed P < 0.05. All statistical analyses were carried out using STATA 13.SE (StataCorp LP, College Station, TX, USA).

### Patient and public involvement

No patients or participants were involved in formulating the research question, establishing the outcome measures, or in the design of the study. No patients or participants were involved in the interpretation of the results or writing of the manuscript. There are no plans to disseminate the results of the research to participants or wider relevant communities.

## 3. Results

Table 1 shows the baseline characteristics of the study population across quartiles of the HRDea-score. The average score was 73 (SD=10), and ranged between 32 and 116. Participants most adherent to the HRD were more likely to be female, have a normal BMI, be highly educated, have never smoked, and consume less calories per day compared to the least adherent.

### HRDea-score and CHD

During a median follow-up of 15.1 years, a total of 2,543 CHD events occurred. High adherence to the HRD was associated with a lower risk of CHD (HR_Q4vsQ1_ 0.85; 95% CI 0.75 – 0.96) in fully adjusted models (Table 2). Adding each potential mediator separately to the multivariable-adjusted model only slightly attenuated the results (Supplementary Table 3). The cumulative effect of all mediators also did not substantially alter the results (HR_Q4vsQ1_ 0.88; 95% CI 0.77 – 0.99).

**Table 3.**
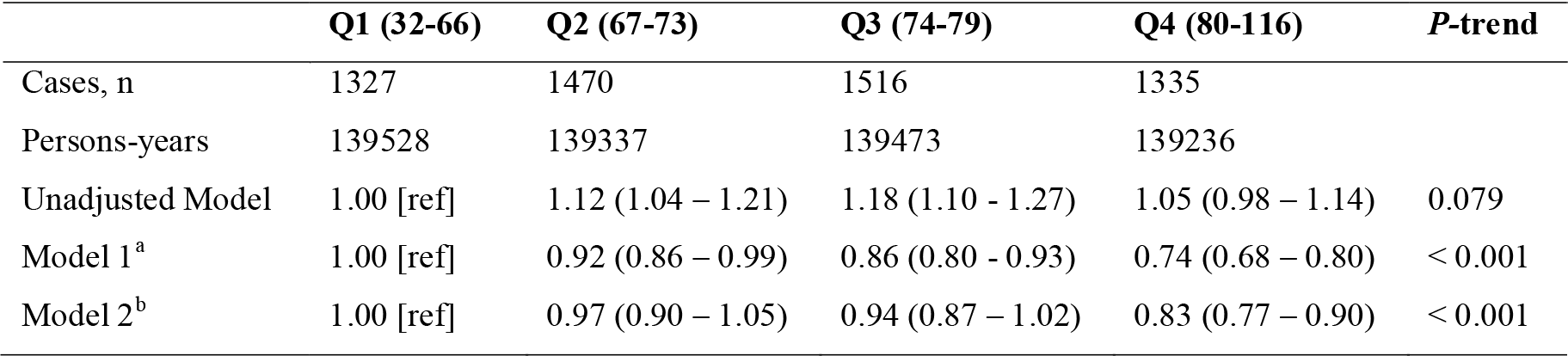
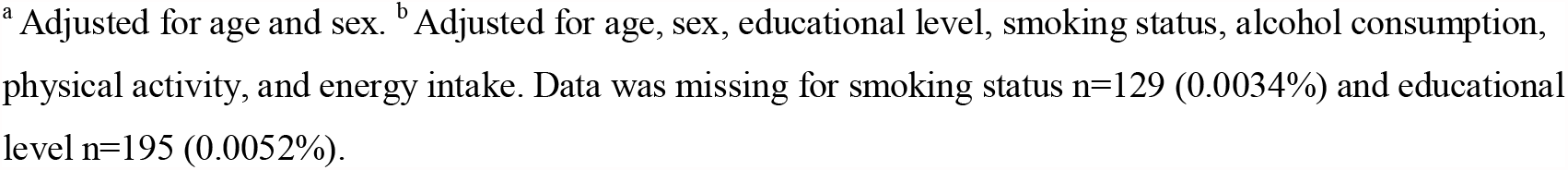
Hazard ratios (HRs) and 95% confidence intervals (CIs) for the association between quartiles of the HRDea-score and all-cause mortality (*n*=37,349).

|  | Q1 (32-66) | Q2 (67-73) | Q3 (74-79) | Q4 (80-116) | P-trend |
| --- | --- | --- | --- | --- | --- |
| Cases, *n* | 1327 | 1470 | 1516 | 1335 |  |
| Persons-years | 139528 | 139337 | 139473 | 139236 |  |
| Unadjusted Model | 1.00 [ref] | 1.12 (1.04 – 1.21) | 1.18 (1.10 - 1.27) | 1.05 (0.98 – 1.14) | 0.079 |
| Model 1<sup>a</sup> | 1.00 [ref] | 0.92 (0.86 – 0.99) | 0.86 (0.80 - 0.93) | 0.74 (0.68 – 0.80) | < 0.001 |
| Model 2<sup>b</sup> | 1.00 [ref] | 0.97 (0.90 – 1.05) | 0.94 (0.87 – 1.02) | 0.83 (0.77 – 0.90) | < 0.001 |

<sup>a</sup> Adjusted for age and sex. <sup>b</sup> Adjusted for age, sex, educational level, smoking status, alcohol consumption, physical activity, and energy intake. Data was missing for smoking status n=129 (0.0034%) and educational level n=195 (0.0052%).

### HRDea-score and all-cause mortality

During a median follow-up of 15.3 years, 5648 people died from all causes. High adherence to the HRD was associated with a lower risk of all-cause mortality (HR_Q4vsQ1_ 0.83; 95% CI 0.77 – 0.90) in fully adjusted models (Table 3).

### HRDea-score and environmental impact

Table 4 shows the baseline means of GHG emissions, land use, blue water use, freshwater eutrophication, marine eutrophication, and terrestrial acidification across quartiles of the HRDea-score. Participants most adherent to the HRD were more likely to consume diets that were associated with less GHGE, land use, freshwater eutrophication, marine eutrophication and terrestrial acidification compared to the least adherent. Yet, diets of those most adherent to the HRD have higher blue water use compared to diets of those least adhering to the HRD.

**Table 4.**
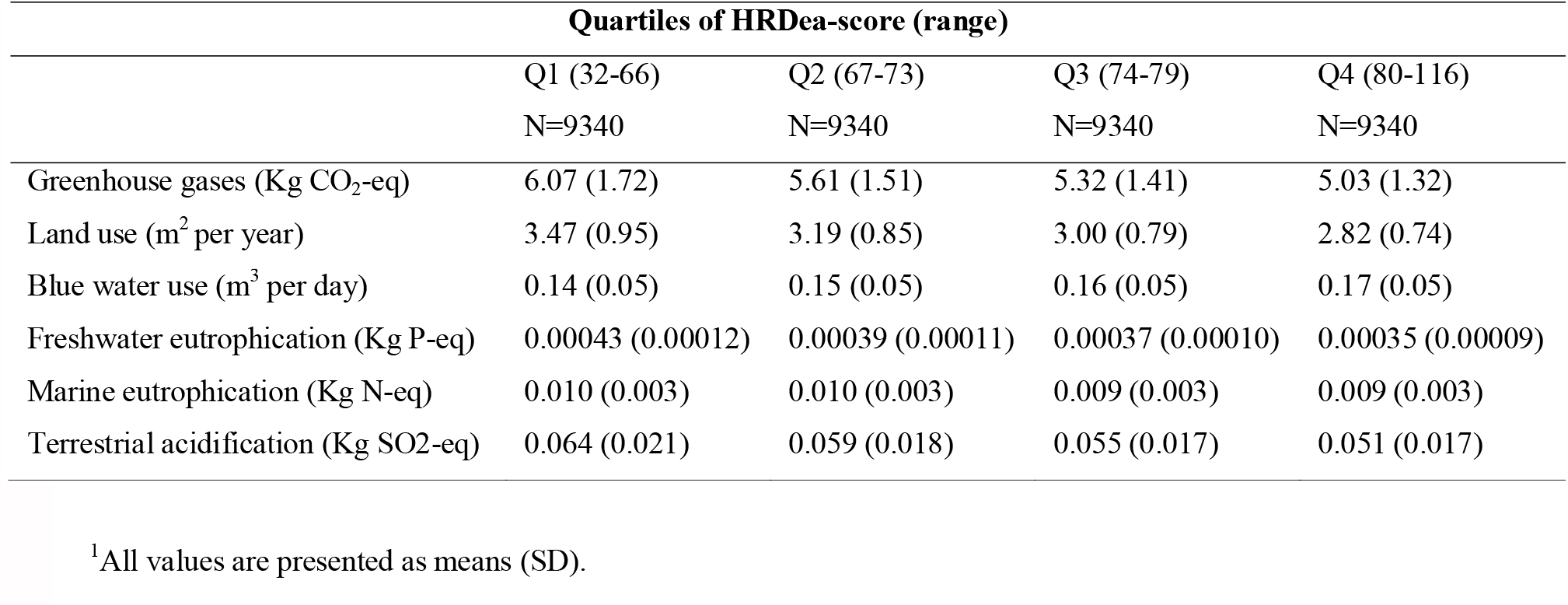
Descriptive statistics of the environmental impact indicators by quartiles of the HRDea-score (*n*=37,393)^1^

In multivariable adjusted models, high adherence to the HRD was associated with lower GHGE (β= -0.10 kg CO2-eq; 95%CI: -0.13, -0.07), less land use (β= -0.11 m^2^ per year; 95% CI: -0.12, -0.09), less freshwater eutrophication (β= -0.000002 kg P-eq; 95%CI: -0.000004, - 0.000001), less marine eutrophication (β= -0.00035 kg N-eq; 95%CI: -0.00042, -0.00029) and less terrestrial acidification (β = -0.004 kg SO2-eq; 95% CI: -0.004, -0.003) and with higher blue water use (β=0.044; 95% CI; 95%CI: 0.043, 0.045) when comparing extreme quartiles (Table 5). These beta-coefficients correspond to 1.7% lower GHGE, 3.2% less land use, 0.5% less freshwater eutrophication, 3.5% less marine eutrophication, 6.3% less terrestrial acidification, but with 31.43% higher blue water use, when comparing extreme quartiles.

**Table 5.** Regression coefficients and 95% confidence intervals (CI) for the association between quartiles of the HRDea-score and environmental indicators (n=37,393).

| HRDea-score (range) |  |  |  |  |  |
| --- | --- | --- | --- | --- | --- |
|  | Q1 (29-67) | Q2 (68-73) | Q3 (74-79) | Q4 (80-114) | P-trend |
| GHGE (kg CO2-eq) |  |  |  |  |  |
| Unadjusted model | 0 [ref] | -0.46 (-0.5, -0.42) | -0.75 (-0.79, -0.71) | -1.04 (-1.08, -0.99) | < 0.001 |
| Model 1<sup>a</sup> | 0 [ref] | -0.05 (-0.07, -0.02) | -0.05 (-0.08, -0.3) | -0.10 (-0.13, -0.07) | < 0.001 |
| Land Use (m<sup>2</sup>/y) |  |  |  |  |  |
| Unadjusted model | 0 [ref] | -0.28 (-0.31, -0.26) | -0.47 (-0.49, -0.45) | -0.66 (-0.68, -0.63) | < 0.001 |
| Model 1<sup>a</sup> | 0 [ref] | -0.04 (-0.06, -0.03) | -0.07 (-0.08, -0.05) | -0.11 (-0.12, -0.09) | < 0.001 |
| Blue water use (m<sup>3</sup>/d) |  |  |  |  |  |
| Unadjusted model | 0 [ref] | 0.01 (0.009, 0.012) | 0.018 (0.017, 0.02) | 0.031 (0.030, 0.032) | < 0.001 |
| Model 1<sup>a</sup> | 0 [ref] | 0.017 (0.016, 0.018) | 0.029 (0.027, 0.03) | 0.044 (0.043, 0.045) | < 0.001 |
| Freshwater eutrophication (Kg P-eq) |  |  |  |  |  |
| Unadjusted model | 0 [ref] | -0.000034 (-0.000036, -0.000031) | -0.000056 (-0.000058, -0.000053) | -0.000074 (-0.000077, -0.000071) | < 0.001 |
| Model 1<sup>a</sup> | 0 [ref] | -0.000002 (-0.000004, -0.000001) | -0.000003 (-0.000004, -0.000001) | -0.000002 (-0.000004, -0.000001) | < 0.001 |
| Marine eutrophication (Kg N-eq) |  |  |  |  |  |
| Unadjusted model | 0 [ref] | -0.0008 (-0.0008, -0.0007) | -0.0013 (-0.0014, -0.0012) | -0.0019 (-0.0020, -0.0018) | < 0.001 |
| Model 1<sup>a</sup> | 0 [ref] | -0.00009 (-0.00015, -0.00002) | -0.00014 (-0.00021, -0.00008) | -0.00035 (-0.00042, -0.00029) | < 0.001 |
| Terrestrial acidification (Kg SO2-eq) |  |  |  |  |  |
| Unadjusted model | 0 [ref] | -0.005 (-0.006, -0.005) | -0.008 (-0.009, -0.008) | -0.012 (-0.013, -0.012) | < 0.001 |
| Model 1<sup>a</sup> | 0 [ref] | -0.001 (-0.001, -0.001) | -0.002 (-0.002, -0.001) | -0.004 (-0.004, -0.003) | < 0.001 |

<sup>a</sup> Adjusted for age, sex, and energy intake.

## Discussion

In the present study among 37,349 Dutch adults, we found that higher adherence to the HRD as proposed by the EAT-Lancet Commission was associated with 15% lower risk of CHD, and with 17% lower risk of all-cause mortality. Higher adherence to the diet was also associated with 1.7% lower GHGE, 3.2% less land use, 0.5% less freshwater eutrophication, 3.5% less marine eutrophication, 6.3% less terrestrial acidification, but with 31.4% higher blue water use.

Before we can interpret our results, we need to address the limitations of the present study. Even though overall the FFQ was considered adequate to assess food intake of the EPIC-NL population, the validity of vegetable and fish intakes was found to be quite poor.^10^ This would suggest possible measurement error in the present study for vegetable and fish intake as well. As misclassification of these food groups is likely to be random given the prospective design of the present study, and considering that these food groups are generally associated with lower risk for CHD, misclassification of the intake of these foods could indicate an attenuation of the inverse association with CHD and mortality risk. Similarly, there may be underestimation of the effects on environmental impact indicators. Moreover, dietary assessment was conducted only at baseline and dietary intake might have changed during follow-up. However, a previous study in EPIC-NL showed dietary changes between baseline and 20 years follow-up to be relatively modest.^43^ Finally, the current study used the Dutch LCA Database to calculate environmental indicators. It should be noted that, although the LCA database is a comprehensive source of LCA indicators, there is also some uncertainty in the data since they are modelled and not actually measured.^41^ Furthermore, LCA estimates for the Netherlands will likely not be fully generalizable to other contexts.

The main strength of this study is the use of a prospective design, based on a large population cohort, and a long follow-up period. Moreover, we used a proportional scoring from 0 to 10 for each component of the HRD-score, which is likely to capture the variability in dietary intake. Additionally, the current study created a refined diet score which could be used or adapted by other studies who wish to study the HRD in other settings. Another strength is the linkage with national registries to ascertain health outcomes which is considered a valid method to reach near-complete follow-up and to reduce possible outcome misclassification.^44^ Finally, the present study included a wide range of environmental indicators, which appeals to the need for an integrated analysis of the core environmental impact dimensions of food systems.^7^

The EAT-Lancet report leaves some space for definition of the HRD, so that recommendations can be tailored to different populations. Thus, for the construction of the HRD-score, several choices were made in assigning foods to each scoring component, such as the inclusion of dairy and starchy vegetables in the optimum component. Depending on the population and cultural context, some might prefer assigning these food groups to an adherence or moderation component. Additionally, intake recommendations in grams per day from the EAT-Lancet report were energy-adjusted for women, to account for their generally lower energy requirements. Since these choices were mostly based on the baseline characteristics of this study population, they might not be entirely appropriate when replicating this study in a different setting.

Findings from the present study are largely in line with the study from Knuppel et al. which used a more simple score to reflect HRD-adherence, and found similar inverse associations for CHD risk and all-cause mortality, although the latter did not reach statistical significance.^8^ Even though the HRD-score is unique to this study, other studies investigating dietary indices focusing on plant-based diets, show inverse associations with either CHD or all-cause mortality.^45 46^ Differences in the magnitude of risk reductions between the present study and available literature are likely related to the scoring methods, the baseline characteristics of the populations, and/or to residual confounding.

With regard to the environmental impact of the HRD, the indicators used in this study are largely in line with the planetary boundaries framework,^38 39^ which is also applied by the EAT-Lancet Commission to model the environmental effects of the HRD. Although there was a significant increase in blue water use, the observed percentage reductions for GHGE, land use, freshwater eutrophication, marine eutrophication and terrestrial acidification in fully adjusted models seem modest. These findings are in line with findings from the EAT-Lancet Commission, showing that dietary changes alone are not sufficient to stay within most planetary boundaries, except for GHGE, for which a reduction of 49% was observed when comparing current diets with the HRD.^7^ The discrepancy in GHGE reductions between the EAT-Lancet report and the current study could be due to the fact that in the EPIC-NL population the maximum HRD-score reached was only 116, while complete adherence would yield 140 points. Thus, observed diets may still be suboptimal, and further improvements towards the HRD may have larger effects on environmental impact indicators. Also, other measures such as improved production practices and less food waste and loss are needed as well.^7^

The Dutch Healthy Diet index 2015 (DHD15-index) – a diet-score reflecting adherence to the Dutch national dietary guidelines – has in EPIC-NL previously been related to environmental sustainability. In line with our findings, these studies also observed adherence to the DHD15 to be associated with lower GHGE and less land use, but with higher use of blue water.^47 48^ Indeed, several plant-based foods – which are emphasized in both the DHD15 and the HRD - do have a relatively high blue water use per kg product, such as several fruits and nuts.^49^ Plant-based foods with a relatively high blue water use are often imported into The Netherlands from areas with a high water scarcity, such as citrus fruits from Spain or almonds from the USA. In order to reduce the blue water footprint of the HRD diet in a Dutch context, choosing locally-grown and seasonal fruits and vegetables may be recommended.^49^ Thus, is plausible that with small changes in the choice for type of fruits, and when choosing for seasonal and locally grown fruits and vegetables, the high blue water use associated with high HRD-adherence could be diminished.

## Conclusion

This study provides evidence from a prospective cohort study in The Netherlands for an inverse association of adherence to the HRD with CHD and all-cause mortality. This research also found that increasing adherence to the HRD could lower some aspects of the environmental impact of diets, but attention is needed for the associated increase in blue water use.

## Data Availability

Data are available upon reasonable request from the PI of the EPIC-NL study (YTvdS).

## Contributor and guarantor information

Contributors: CC, MCH, and YTvdS designed the study. CC performed the statistical analyses. REV and EHMT contributed to the linkage of dietary data with environmental data. CC drafted the manuscript. MCH, REV, WMMV, JMAB, EHMT, YTvdS reviewed and revised the manuscript.

YTvdS is the guarantor. The corresponding author attests that all listed authors meet authorship criteria and that no others meeting the criteria have been omitted.

## Copyright/license for publication

The corresponding author has the right to grant on behalf of all authors and does grant on behalf of all authors, a worldwide licence to the Publishers and its licensees in perpetuity, in all forms, formats and media (whether known now or created in the future), to i) publish, reproduce, distribute, display and store the Contribution, ii) translate the Contribution into other languages, create adaptations, reprints, include within collections and create summaries, extracts and/or, abstracts of the Contribution, iii) create any other derivative work(s) based on the Contribution, iv) to exploit all subsidiary rights in the Contribution, v) the inclusion of electronic links from the Contribution to third party material where-ever it may be located; and, vi) licence any third party to do any or all of the above.

## Data availability statement

Data are available upon reasonable request from the PI of the EPIC-NL study (YTvdS).

## Ethical approval

The study complied with the Declaration of Helsinki and was approved by the Institutional Review Board of the University Medical Center Utrecht (Prospect) and the Medical Ethical Committee of the Netherlands Organization for Applied Scientific Research (TNO) (MORGEN). All participants proved written informed consent.

## Transparency statement

YTvdS affirms that the manuscript is an honest, accurate, and transparent account of the study being reported; that no important aspects of the study have been omitted; and that any discrepancies from the study as originally planned (and, if relevant, registered) have been explained.

## Funding

MCH was supported by the Netherlands Cardiovascular Research Initiative, an initiative with support of the Dutch Heart Foundation (CVON2016-04) and The Netherlands Organisation for Health Research and Development (531003001) in the context of the Supreme Nudge project. Funders had no role in the design of the study and collection, analysis, and interpretation of data and in writing the manuscript, nor have they authority on the decision to submit the manuscript for publication.

### Section 1: What is already known on this topic

- The EAT-Lancet Healthy Reference Diet (HRD) was suggested to co-benefit public and planetary health.
- Only one study empirically assessed the association between the HRD and the risk of coronary heart disease and all-cause mortality, showing that better adherence to the HRD was associated with 28% lower risk for coronary heart disease, but not with risk of all-cause mortality.
- No study has yet empirically evaluated the environmental impact of the HRD.

### Section 2: What this study adds

- The present study suggests that adhering to the HRD is associated with lower risk of coronary heart disease and all-cause mortality, and is associated with lower environmental impact for the majority of environmental impact indictors studied.
- Adherence to the HRD was, however, associated with increase blue water use, which warrants further attention.

