## Supplementary Material for "Adherence to the EAT-Lancet Healthy Reference Diet in relation to Coronary Heart Disease, All-Cause Mortality Risk and Environmental Impact: Results from the EPIC-NL Cohort"

40,011

Baseline EPIC-NL cohort (PROSPECT n= 17,357; MORGEN n=22,654)

38,345

Individuals who withdrew permission for linkage with disease registries (n=1,666)

37,350

Individuals with extreme energy intake (n=400)

37,568

Prevalent cases of CHD (n=377)

37,945

Individuals with incomplete FFQ (n=218)

37,349

Individuals who withdrew consent (n=1)

37,349

Final study population

**Supplementary Figure 1.** Flowchart of the study population.

**Supplementary Table 1.** Construction of HRD-score, intake recommendations (g/day) and scoring system for men based on 2500 kcal/day.

| **Food Group** | **Component type^1^** | **HRD recommendation (g/day)** | **Minimum score**  **(0 points)** | **Proportional score** | **Maximum points**  **(10 points)** | **Proportional score** |
| --- | --- | --- | --- | --- | --- | --- |
| **Whole Grains^2^** |  |  |  |  |  |  |
| Rice, wheat, corn, and other | A | 464 (or 60% of total energy) | 0 g/d | 0-464 g/d | ≥464 g/d |  |
| **Vegetables** |  |  |  |  |  |  |
| All vegetables^3^ | A | 300 | 0 g/d | 0-300 g/d | ≥300 g/d |  |
| **Fruits** |  |  |  |  |  |  |
| All fruit^4^ | A | 200 | 0 g/d | 0-200 g/d | ≥200 g/d |  |
| **Tubers or starchy vegetables** |  |  |  |  |  |  |
| Potatoes and cassava | O | 50 | 0 g/d | 0-50 g/d | 50-100 g/d | 100-150 g/d |
| **Dairy foods** |  |  |  |  |  |  |
| Whole milk or derivative equivalents (e.g., cheese) | O | 250 | 0 g/d | 0-250 g/d | 250 – 500 g/d | 500-750 g/d |
| **Protein Sources** |  |  |  |  |  |  |
| Dry beans, lentils, and peas | A | 50 | 0 g/d | 0-50 g/d | ≥50 g/d |  |
| Soy foods | A | 25 | 0 g/d | 0-25 g/d | ≥25 g/d |  |
| Beef, lamb and pork | M | 14 | ≥14 g/d | 14 -0 g/d | 0 g/d |  |
| Chicken and other poultry | O | 29 | 0 g/d | 0-29 g/d | 29-58 g/d | 58-88 g/d |
| Eggs | O | 13 | 0 g/d | 0-13 g/d | 13-25 g/d | 25-38 g/d |
| Fish | O | 28 | 0 g/d | 0-28 g/d | 28-100 g/d | 100-128 g/d |
| Nuts | O | 50 | 0 g/d | 0-50 g/d | 50-100 g/d | 100-150 g/d |
| **Added sugars** |  |  |  |  |  |  |
| All sweeteners | M | 31 | ≥31 g/d | 31-0 g/d | 0 g/d |  |
| **Added fats** | R |  |  |  |  |  |
| Palm oil |  | 6.8 | No consumption of unsaturated fats OR ratio of unsaturated to saturated fats of ≤0.6^5^ |  | No consumption of saturated fats OR ratio of unsaturated to saturated fats of ≥13^5^ |  |
| unsaturated oils |  | 40 |  |  |  |  |
| dairy fats included in milk |  | 0 |  |  |  |  |
| lard and tallow |  | 5 |  |  |  |  |

^1^A = adequacy component; O = optimum component; M = moderation component; R = ratio component.

^2^ Reference diet refers to dry, raw weight. Recommendations for whole grains were converted, as described by Dooren et al.^1^

^3^ Including dark green vegetables, red and orange vegetables, other vegetables.

^4^ Excluding fruit juice.

^5^ Cut-offs and threshold values were derived from the 15th percentile and 85th percentile of the intake distribution of the Dutch reference population, as described in Looman et al.^2^

**Supplementary Table 2.** Construction of HRD-score, intake recommendations (g/day) and scoring system for women based on 2000 kcal/day.

| **Food Group** | **Component type^1^** | **HRD recommendation (g/day)** | **Minimum score**  **(0 points)** | **Proportional score** | **Maximum points**  **(10 points)** | **Proportional score** |
| --- | --- | --- | --- | --- | --- | --- |
| **Whole Grains^2^** |  |  |  |  |  |  |
| Rice, wheat, corn, and other | A | 372 (or 60%en) | 0 g/d | 0-372 g/d | ≥372 g/d |  |
| **Vegetables** |  |  |  |  |  |  |
| All vegetables^3^ | A | 240 | 0 g/d | 0-240 g/d | ≥240 g/d |  |
| **Fruits** |  |  |  |  |  |  |
| All fruit^4^ | A | 160 | 0 g/d | 0-160 g/d | ≥160 g/d |  |
| **Tubers or starchy vegetables** |  |  |  |  |  |  |
| Potatoes and cassava | O | 40 | 0 g/d | 0-40 g/d | 40-80 g/d | 80-120 g/d |
| **Dairy foods** |  |  |  |  |  |  |
| Whole milk or derivative equivalents (e.g., cheese) | O | 200 | 0 g/d | 0-200 g/d | 200 – 400 g/d | 400-600 g/d |
| **Protein Sources** |  |  |  |  |  |  |
| Dry beans, lentils, and peas | A | 40 | 0 g/d | 0-40 g/d | ≥40 g/d |  |
| Soy foods | A | 20 | 0 g/d | 0-20 g/d | ≥20 g/d |  |
| Beef, lamb and pork | M | 12 | ≥12 g/d | 12-0 g/d | 0 g/d |  |
| Chicken and other poultry | O | 23 | 0 g/d | 0-23 g/d | 23-46 g/d | 46-69 g/d |
| Eggs | O | 10 | 0 g/d | 0-10 g/d | 10-20 g/d | 20-30 g/d |
| Fish | O | 22 | 0 g/d | 0-22 g/d | 22-80 g/d | 80-102 g/d |
| Nuts | O | 40 | 0 g/d | 0-40 g/d | 40-80 g/d | 80-120 g/d |
| **Added sugars** |  |  |  |  |  |  |
| All sweeteners | M | 25 | ≥25 g/d | 25-0 g/d | 0 g/d |  |
| **Added fats** | R |  |  |  |  |  |
| Palm oil |  | 5 | No consumption of unsaturated fats OR ratio of unsaturated to saturated fats of ≤0.5^5^ |  | No consumption of unsaturated fats OR ratio of unsaturated to saturated fats of ≤0.5^5^ |  |
| unsaturated oils |  | 32 |  |  |  |  |
| dairy fats included in milk |  | 0 |  |  |  |  |
| lard and tallow |  | 4 |  |  |  |  |

^1^A = adequacy component; O = optimum component; M = moderation component; R = ratio component.

^2^ Reference diet refers to dry, raw weight. Recommendations for whole grains were converted, as described by Dooren et al.^1^

^3^ Including dark green vegetables, red and orange vegetables, other vegetables.

^4^ Excluding fruit juice.

^5^ Cut-offs and threshold values were derived from the 15th percentile and 85th percentile of the intake distribution of the Dutch reference population, as described in Looman et al.^2^

**Supplementary Table 3.** Hazard ratios (HR) and 95% confidence intervals (CI) for the association between quartiles of the HRDea-score and CHD, with adjustment for mediating factors (*n*=37,349).

|  | **HRDea-score (range)** | | | |  |
| --- | --- | --- | --- | --- | --- |
|  | **Q1 (32-66)** | **Q2 (67-73)** | **Q3 (74-79)** | **Q4 (80-116)** | ***P*-trend** |
| Cases, *n* (*n* total*=*2543) | 700 | 684 | 606 | 553 |  |
| HR (95% CI) |  |  |  |  |  |
| Model 2 + BMI | 1.00 [ref] | 0.98 (0.88 – 1.10) | 0.88 (0.79 – 0.99) | 0.88 (0.78 – 1.00) | 0.019 |
| Model 2 + cholesterol | 1.00 [ref] | 0.96 (0.86 – 1.07) | 0.85 (0.76 – 0.96) | 0.85 (0.75 – 0.96) | 0.003 |
| Model 2 + hypertension | 1.00 [ref] | 0.96 (0.87 -1.07) | 0.86 (0.77 – 0.97) | 0.87 (0.77 – 0.98) | 0.006 |
| Model 2 + all possible mediators | 1.00 [ref] | 0.98 (0.87 – 1.09) | 0.87 (0.78 – 0.98) | 0.88 (0.77 – 0.99) | 0.014 |
